# Phenotypic and genetic associations between anhedonia and brain structure in UK Biobank

**DOI:** 10.1101/2020.06.29.20142984

**Authors:** Xingxing Zhu, Joey Ward, Breda Cullen, Donald M. Lyall, Rona J. Strawbridge, Daniel J. Smith, Laura M. Lyall

## Abstract

**Background:** Anhedonia is a core symptom of multiple psychiatric disorders and has been associated with changes in brain structure. Genome-wide association studies suggest that anhedonia is heritable with a polygenic architecture but few studies have explored the association between genetic loading for anhedonia - indexed by polygenic risk scores for anhedonia (PRS-anhedonia) - and structural brain imaging phenotypes. We investigated how anhedonia and polygenic risk for anhedonia were associated with brain structure within the UK Biobank cohort.

**Methods:** Brain measures (including total grey/white matter volumes, subcortical volumes, cortical thickness and white matter integrity) were analysed in relation to the self-reported anhedonia phenotype and PRS-anhedonia for 17,492 participants (8,506 males and 8,986 females; mean age = 62.81 years, SD = 7.43), using linear mixed models and including mediation analyses.

**Results:** State anhedonia was significantly associated with smaller total grey matter volume (GMV), smaller volumes in thalamus and nucleus accumbens; as well as reduced cortical thickness within the paracentral gyrus, the opercular part of inferior frontal gyrus and the rostral anterior cingulate cortex. PRS-anhedonia was associated with reduced total GMV, increased total white matter volume and reduced white matter integrity; in addition to reduced cortical thickness within the parahippocampal cortex, the superior temporal gyrus and the insula cortex.

**Conclusions:** Both the state anhedonia phenotype and PRS-anhedonia were associated with differences in multiple brain structures/areas, including within reward-related circuits. These differences may represent vulnerability markers for psychopathology across a range of psychiatric disorders.

## Introduction

Anhedonia, defined as subjectively diminished capacity to experience pleasure, is a trans-diagnostic symptom present in several psychiatric disorders, including major depressive disorder (MDD) and schizophrenia (1, 2). In recent years, with the development of the Research Domain Criteria (RDoC) initiative (3), research efforts focused on trans-diagnostic symptoms such as anhedonia have grown substantially, including progress on describing the underlying genetic architecture of anhedonia (4) and brain regions associated with anhedonia (e.g., 5). As a symptom that cuts across traditional diagnostic categories, anhedonia represents a promising RDoC construct with potential to elucidate some of the underlying biology of psychiatric disorders.

Anhedonia is associated with impairments in reward processing and may be focused on frontal-striatal brain circuits (6-8). Evidence from neuroimaging studies suggests that alterations in reward-related regions may contribute to the subjective experience of anhedonia. For example, studies in clinical populations (particularly MDD and schizophrenia), report reductions in grey matter volume (GMV) and cortical thickness (CT) within several brain regions, including the caudate, the nucleus accumbens (NAcc), the anterior cingulate cortex (ACC), the prefrontal cortex and the parietal lobe (9-12). Similarly, within non-clinical populations, anhedonia may be associated with reduced volume in the caudate (13) and the NAcc (14, 15), although these findings have been inconsistent (in healthy controls Lee *et al*. (12) reported that anhedonia was associated with increased GMV in the posterior cingulate cortex and precuneus and, in subjects with MDD, Yang *et al*. (16) found no correlation between anhedonia and both prefrontal CT or parietal CT). Such findings may reflect relatively small sample sizes (clinical sample N<100 (e.g., 10, 11, 12, 16) and non-clinical sample N≤ 50 (e.g., 13, 14, 15)) and both demographic and clinical heterogeneity within samples.

Anhedonia may also be associated with abnormalities of structural connectivity between brain regions although findings have not reached consistency. A study of negative symptoms in schizophrenia reported negative associations between anhedonia and fractional anisotropy (FA) of left frontal lobe (17), while another study found anhedonia was positively correlated with FA in the cingulum and superior longitudinal fasciculus (SLF) in patients with schizophrenia (18). In addition, Coloigner *et al*. (19) found in depressed patients higher <u>anhedonia</u> was correlated with greater FA in the SLF and lower FA in the cingulum, the genu of <u>corpus callosum</u> and the posterior thalamic radiation. Further, Yang *et al*. (20) found that, in healthy controls, anhedonia was associated with higher mean FA in the SLF, the anterior thalamic radiation (ATR) and the corticospinal tract. Regardless of the directional inconsistency, abnormalities in these structures tentatively support the assertion that anhedonia involves abnormal and/or inefficient communication between brain regions, especially structures within reward circuits, which might in turn contribute to vulnerability to psychiatric disorders, such as MDD (6, 7). It is notable that these studies of white matter integrity were also conducted with small numbers of participants (e.g., less than fifty patients or healthy controls in 18, 20) and most studies to date on anhedonia and brain structure have been conducted in highly selected clinical samples of MDD and schizophrenia.

In contrast to recent large-scale genome-wide association studies (GWAS) of MDD and schizophrenia (21, 22), most GWAS of anhedonia have been small and underpowered. A meta-analysis of three GWAS studies (total sample size 6,297) reported just a single locus associated with anhedonia in the discovery sample and no replication (23). More recently, a large GWAS (*n*=375,275) of state anhedonia identified 11 novel loci (4). This study also found evidence for significant genetic correlations between anhedonia and both MDD and schizophrenia (4, 23). So far, only a small number of imaging studies have examined the association between polygenic risk for anhedonia and brain structure (4, 24). In preliminary analyses, Lyall and Ward *et al*. (4) found associations between genetic predisposition to anhedonia and smaller total GMV, smaller volume of the orbitofrontal cortex, middle frontal gyrus and insula, in addition to worse brain white matter integrity (lower FA; higher MD).

Our goal was to make use of the extensive data within the UK Biobank cohort, including the most recent release of neuroimaging data with greater numbers and a broader range of informative structural phenotypes, to: a) test for associations between state anhedonia and brain structure; b) test for association between PRS-anhedonia and brain structure (including subcortical volumes, white matter integrity and CT); and c) to conduct mediation analyses to assess whether brain structure or manifestation of anhedonia/depression might represent intermediates of PRS-anhedonia.

## Methods and Materials

### Participants

The sample included in this study was drawn from UK Biobank, which gathered extensive questionnaire, physical and cognitive measures, as well as biological samples from over 500,000 participants. Informed consent was obtained from all participants, and this study was conducted under approval from the NHS National Research Ethics Service (UK Biobank approved applications #6553 and #17689).

At the first imaging visit, a total of 31,064 participants responded to the question about anhedonia (data field 2060). Participants were excluded for the following reasons: having a developmental, neurological or mental disorder (See Table S1 for detailed participant exclusion criteria); not of White European ancestry; unclear values of anhedonia (responded as ‘prefer not to answer’ or ‘do not know’); age at magnetic resonance imaging (MRI) result missing; brain imaging data unavailable; and outliers (± 3SDs) for total brain volume (sum of total grey matter, white matter and ventricular cerebrospinal fluid volume). A total of 17,492 participants (ages 45-80 years, 8,506 males) were included in this study (see Table 1 and Fig S1). For PRS analyses, participants included in the GWAS of anhedonia were also excluded to avoid overlapping between the train and test samples.

**Table 1.**
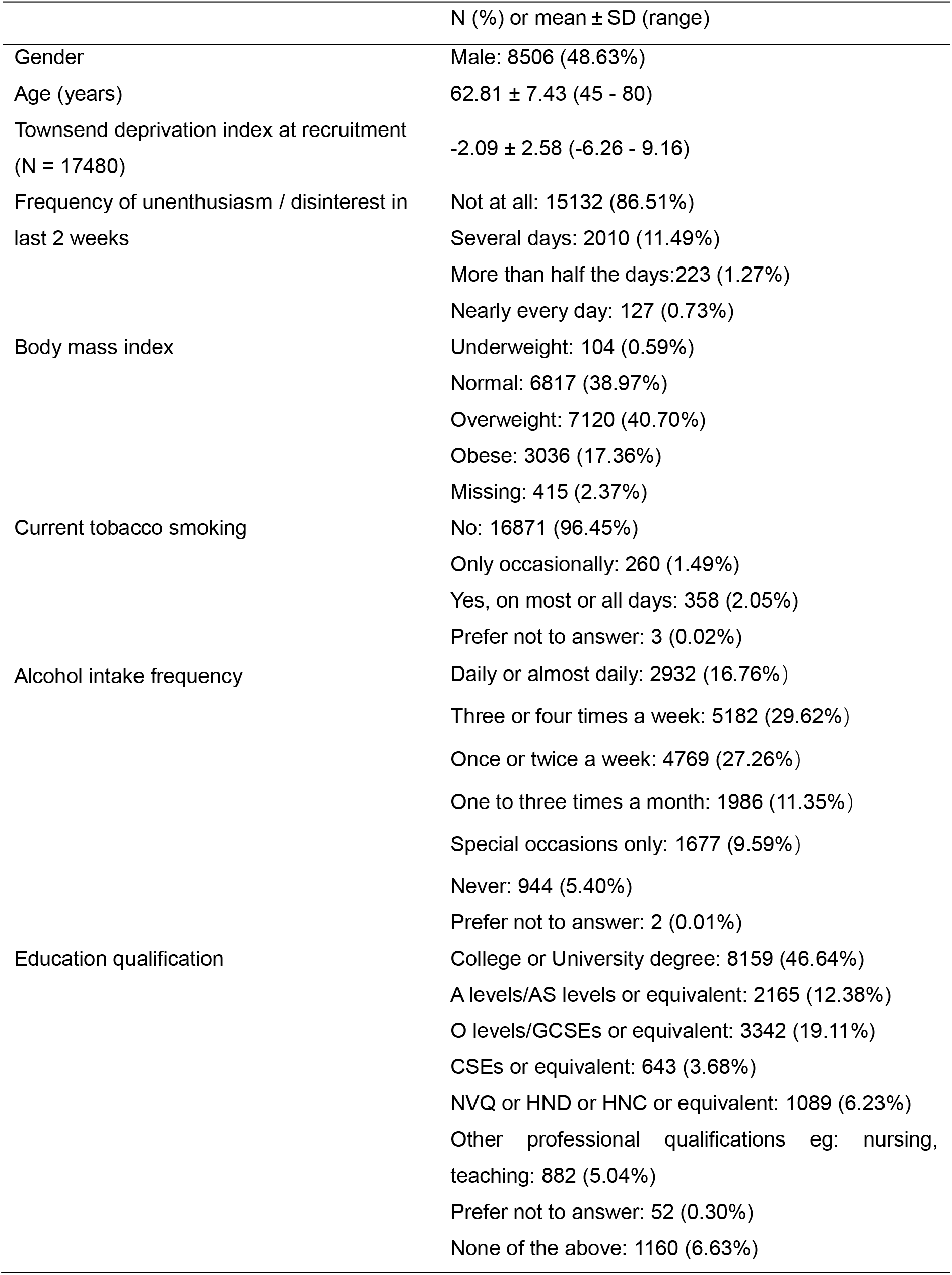
Descriptive statistics for demographic variables (N=17492). The descriptive statistics below are summarized based on the sample included in analyses for anhedonia.

| | N (%) or mean ± SD (range) |
| --- | --- |
| Gender | Male: 8506 (48.63%) |
| Age (years) | 62.81 ± 7.43 (45 - 80) |
| Townsend deprivation index at recruitment<br>(N = 17480) | -2.09 ± 2.58 (-6.26 - 9.16) |
| Frequency of unenthusiasm / disinterest in<br>last 2 weeks | Not at all: 15132 (86.51%)<br>Several days: 2010 (11.49%)<br>More than half the days: 223 (1.27%)<br>Nearly every day: 127 (0.73%) |
| Body mass index | Underweight: 104 (0.59%)<br>Normal: 6817 (38.97%)<br>Overweight: 7120 (40.70%)<br>Obese: 3036 (17.36%)<br>Missing: 415 (2.37%) |
| Current tobacco smoking | No: 16871 (96.45%)<br>Only occasionally: 260 (1.49%)<br>Yes, on most or all days: 358 (2.05%)<br>Prefer not to answer: 3 (0.02%) |
| Alcohol intake frequency | Daily or almost daily: 2932 (16.76%)<br>Three or four times a week: 5182 (29.62%)<br>Once or twice a week: 4769 (27.26%)<br>One to three times a month: 1986 (11.35%)<br>Special occasions only: 1677 (9.59%)<br>Never: 944 (5.40%)<br>Prefer not to answer: 2 (0.01%) |
| Education qualification | College or University degree: 8159 (46.64%)<br>A levels/AS levels or equivalent: 2165 (12.38%)<br>O levels/GCSEs or equivalent: 3342 (19.11%)<br>CSEs or equivalent: 643 (3.68%)<br>NVQ or HND or HNC or equivalent: 1089 (6.23%)<br>Other professional qualifications eg: nursing,<br>teaching: 882 (5.04%)<br>Prefer not to answer: 52 (0.30%)<br>None of the above: 1160 (6.63%) |

### State anhedonia phenotype

As previously described (4), state anhedonia was assessed by a single question, *“Over the past two weeks, how often have you had little interest or pleasure in doing things?”*. Participants could choose from the following answers: “not at all”; “several days”; “more than half the days”; and “nearly every day”, which were coded as 0, 1, 2 and 3 respectively.

### Brain imaging variables

All neuroimaging data were acquired, pre-processed, quality controlled and made available by UK Biobank (https://biobank.ctsu.ox.ac.uk/crystal/crystal/docs/brain_mri.pdf). Details on the acquisition parameters (http://biobank.ctsu.ox.ac.uk/crystal/refer.cgi?id=1977) and the imaging protocol (http://biobank.ctsu.ox.ac.uk/crystal/refer.cgi?id=2367) are documented online and are described within protocol papers (25). The neuroimaging data analysed in the current study consisted of: (1) total GMV and total white matter volume (WMV); (2) subcortical volumes; (3) CT of 31 regions in each hemisphere from the “Desikan–Killiany–Tourville” (DKT) protocol (26) over whole brain surface; and (4) white matter microstructure, indexed by fractional anisotropy (FA) and mean diffusivity (MD). A more detailed description of these variables is provided within supplementary materials.

### Derivation of polygenic risk scores for anhedonia

Polygenic risk scores were calculated using LDpred (27) applied to the summary statistics from a recent GWAS of state anhedonia (4). Of note, the individuals with neuroimaging data in the current study were excluded from this GWAS (4). Additional exclusion criteria for participants in the current study included: over 10% of genetic data missing; self-reported sex did not match genetic sex; purported sex chromosome aneuploidy was reported; where the heterozygosity value was a clear outlier; and participants were not of White European ancestry. In total, 14,904 participants were included in PRS-anhedonia analyses (see supplementary Table S2 and Fig S1).

### Statistical analysis

Data analysis was conducted using Stata. All brain measures were rescaled into zero mean and unitary standard deviation so that the effect sizes represent standardized scores. For each MRI outcome, as noted above, data points with values more than 3 standard deviations from the sample mean were iteratively excluded. False Discovery Rate (FDR) correction at a rate of p<0.05 was applied across all 40 volume/thickness measures and 30 white matter integrity indexes using ‘p.adjust’ function in R (28, 29).

For associations between state anhedonia and brain structures, anhedonia was set as an independent predictor, and each neuroimaging measure was set as a dependent variable. For bilateral brain measures, anhedonia × hemisphere interactions were firstly examined in a repeated measures format to determine whether analysis of left and right homologous structures separately was required, with sex, age, age^2^, hemisphere, ICV and scanner positions on the x, y and z axes set as covariates. Where there was a significant anhedonia × hemisphere interaction, analyses on both lateralised structures were conducted additionally. For whole-brain or single structures, a general linear model was applied without controlling for hemisphere. In the main analysis, the model included sex, age, age^2^, total ICV, scanner positions on the x, y and z axes and hemisphere as fixed effects using repeated measure design. For the PRS-anhedonia analyses, PRS-anhedonia was set as a predictor and genotype array and the first ten genetic principal components were added, in addition to the above covariates, for all association tests.

Furthermore, we conducted sensitivity analyses to assess the robustness of any observed associations with brain structures. Firstly, we set anhedonia as a dichotomous variable, with value ‘0’ referring to participants who reported ‘not at all’, and a value of ‘1’ representing the remainder of participants (who had reported any anhedonia), and re-analysed its relationship with brain measures associated with anhedonia using same models. Secondly, we assessed whether brain differences associated with state anhedonia in this healthy group (those without psychiatric disorder) might represent vulnerability markers for depression. We selected a separate group of participants with depression/post-natal depression (see Table S3 for participant information) according to self-reported psychological and psychiatric problems (data field 20002) and tested for an interaction between anhedonia and depression/post-natal depression on imaging measures in a combined sample with healthy participants and those with depression. Thirdly, we tested whether brain measures associated with state anhedonia or PRS-anhedonia remain significant when Townsend social deprivation index, education qualification, body mass index (BMI), current tobacco use and alcohol intake frequency were set as additional covariates. Each of these covariates is described in detail in supplementary materials.

Finally, in order to explore possible mediation candidates of PRS-anhedonia, we examined the relationship between anhedonia, PRS-anhedonia and brain measures associated with anhedonia or its polygenic risk using mediation analyses. Although we could not determine the directional or causal relationship of these associations, we tested whether brain structures might act as mediators between PRS-anhedonia and state anhedonia and whether state anhedonia could mediate the effect of PRS-anhedonia on brain structures. All covariates remained the same as in the analyses for associations between PRS-anhedonia and brain structure. Mediation analysis were conducted using the ‘sem’ command in Stata and the significance of the mediation effect was estimated using the bootstrap approach (with 1000 random samplings). For bilateral brain structures, values from the left and right hemispheres were averaged for each participant given that the interaction between anhedonia and hemisphere was not significant. FDR correction was applied for two types of mediation models respectively.

## Results

### Demographics

State anhedonia was significantly negatively correlated with age (Pearson’s r = −0.097, p < 0.001), and positively correlated with Townsend social deprivation index (Pearson’s r = 0.068, p < 0.001). Independent t-test showed there was no significant sex difference for anhedonia (t = −0.582, p = 0.560). In addition, One-way ANOVA results showed significant difference in anhedonia between different groups of BMI (F = 33.59, p < 0.001), current tobacco smoking status (F = 23.53, p < 0.001), alcohol intake frequency (F = 13.33, p < 0.001), and education qualification (F = 12.83, p < 0.001). PRS-anhedonia significantly predicted state anhedonia (β = 0.027, F_(1,14902)_ = 53.11, R-squared = 0.004, p < 0.001). We also conducted these analyses using non-parametric tests and the results remained the same (see supplemental results).

### Associations between state anhedonia and brain morphometric measures

For whole-brain measures, we found that state anhedonia was associated with reduced total GMV (β = − 0.018, p_corrected_ = 0.010; Table 2). For bilateral subcortical and cortical measures, no region demonstrated significant interaction of hemisphere, therefore no region was examined separately on different hemispheres (Table S4). Analyses using the repeat measure design showed there were significant associations between state anhedonia and smaller volume of the thalamus (β = −0.040, p_corrected_ < 0.001) and NAcc (β = −0.050, p_corrected_ < 0.001), and with reduced CT in the paracentral gyrus (β = −0.044, p_corrected_ = 0.024; see Fig. 1A for cortical associations), rostral ACC (β = −0.044, p_corrected_ = 0.010) and opercular part of inferior frontal gyrus (pars opercularis; β = −0.040, p_corrected_ = 0.040).

**Table 2.**
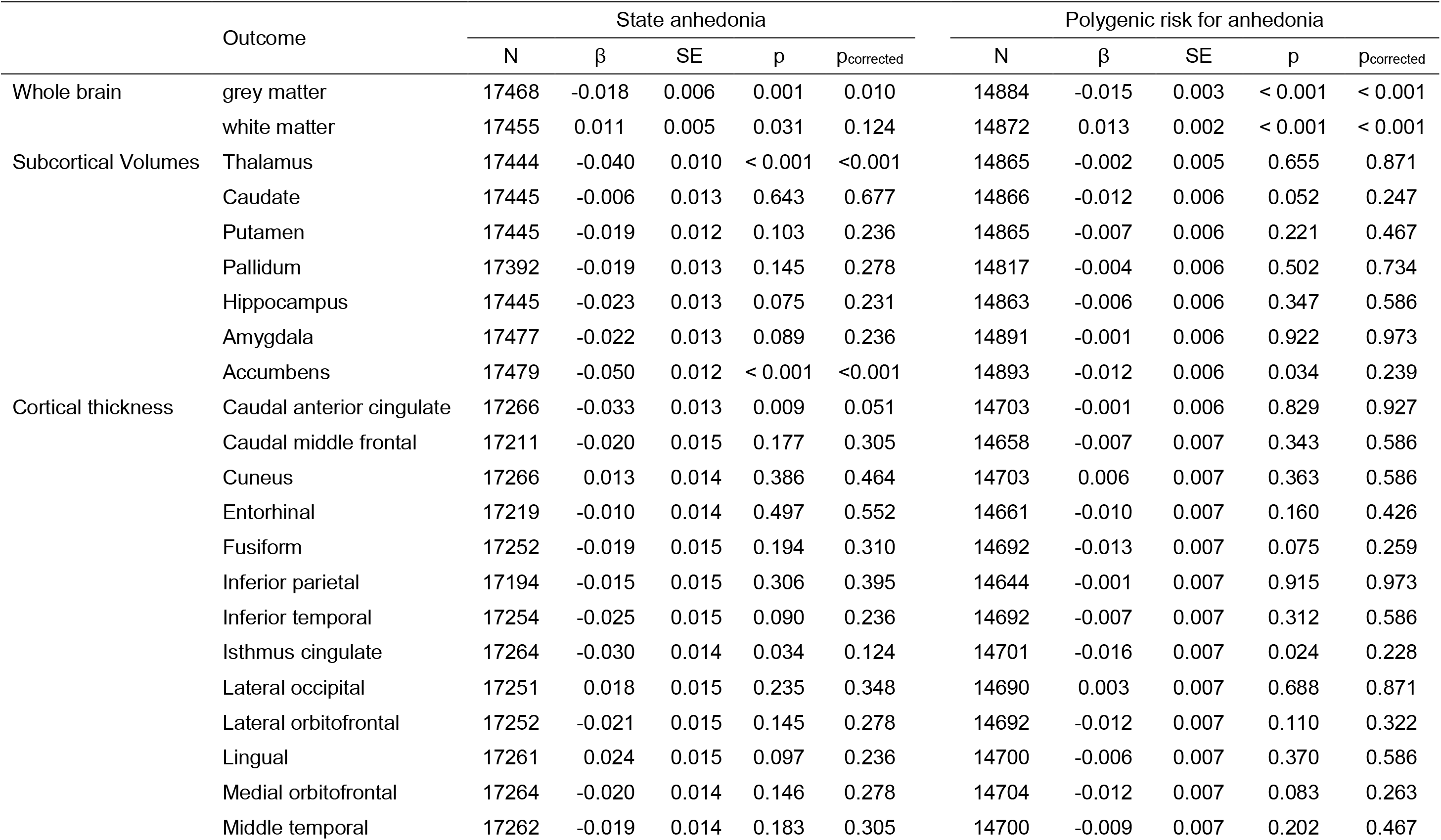
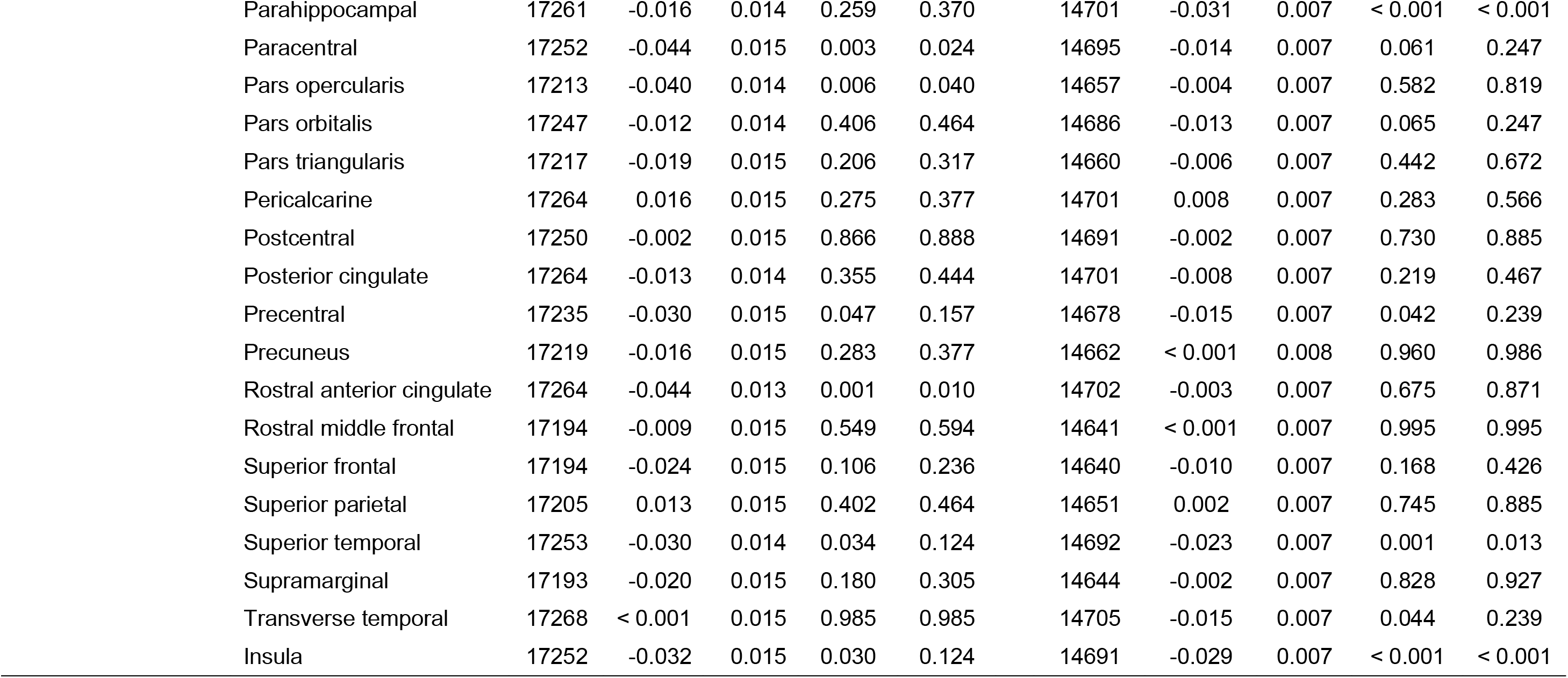
The associations between anhedonia, PRS-anhedonia, brain volumes and cortical thickness.

**Figure 1:**
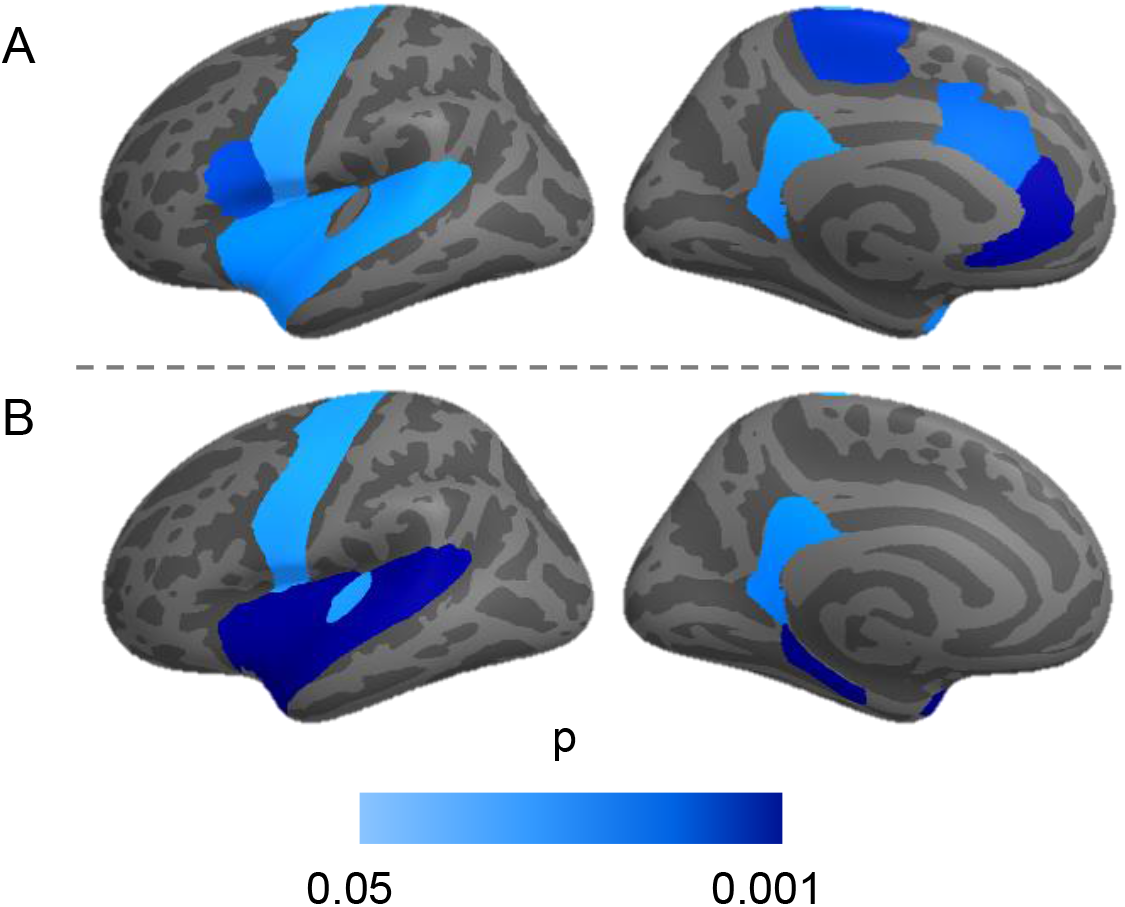
Cortical map of associations of anhedonia, polygenic risk score and cortical thickness. (A) Associations between anhedonia and cortical thickness; (B) Regions associated with polygenic risk score for anhedonia. Regions with uncorrected threshold between p = 0.05 and p = 0.001 were mapped on the left hemisphere.

**Fig 2.**
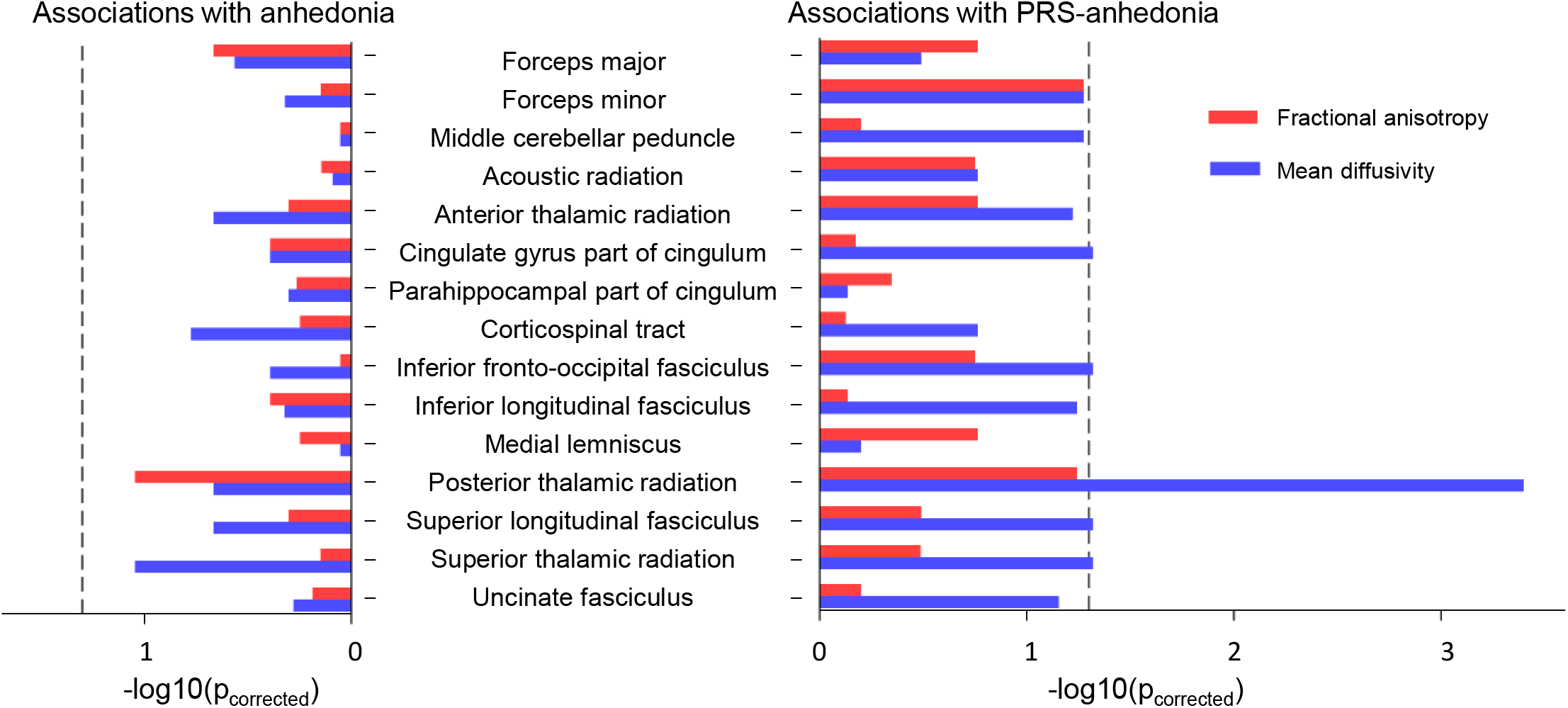
Associations between anhedonia and polygenic risk for anhedonia and white matter integrity. The left panel shows none of the white matter tracts was significantly associated with anhedonia after correction. The right panel shows mean diffusivity in several tracts was significantly associated with polygenic risk for anhedonia. The gray dashed line indicate a significant level (*p* = 0.05). The p value for posterior thalamic radiation was assumed to be 0.0004 when drawing this bar plot because the reported p value from the regression analysis was less than 0.0001.

### Associations between anhedonia and measures of white matter integrity

The same models were applied to analyses of white matter tracts (FA and MD). We firstly tested the interaction of hemisphere and state anhedonia and found significant effects on FA of the uncinate fasciculus (β = −0.056, p_corrected_ < 0.001; Table S5), MD of the acoustic radiation (β = −0.055, p_corrected_ = 0.016) and MD of the uncinate fasciculus (β = 0.047, p_corrected_ < 0.001). Therefore, the left and right FA and MD values of these tracts were also tested (Table 3). Although FA in the forceps major (β = −0.035, p_uncorrected_ = 0.042; Table 3), and MD in the corticospinal tract (β = 0.039, p_uncorrected_ = 0.014), SLF (β = 0.033, p_uncorrected_ = 0.040), ATR (β = 0.034, p_uncorrected_ = 0.024), superior thalamic radiation (β = 0.044, p_uncorrected_ = 0.003) and posterior thalamic radiation (β = 0.015, p_uncorrected_ = 0.030) showed a possible pattern of association, none of these associations survived correction for multiple testing.

**Table 3.**
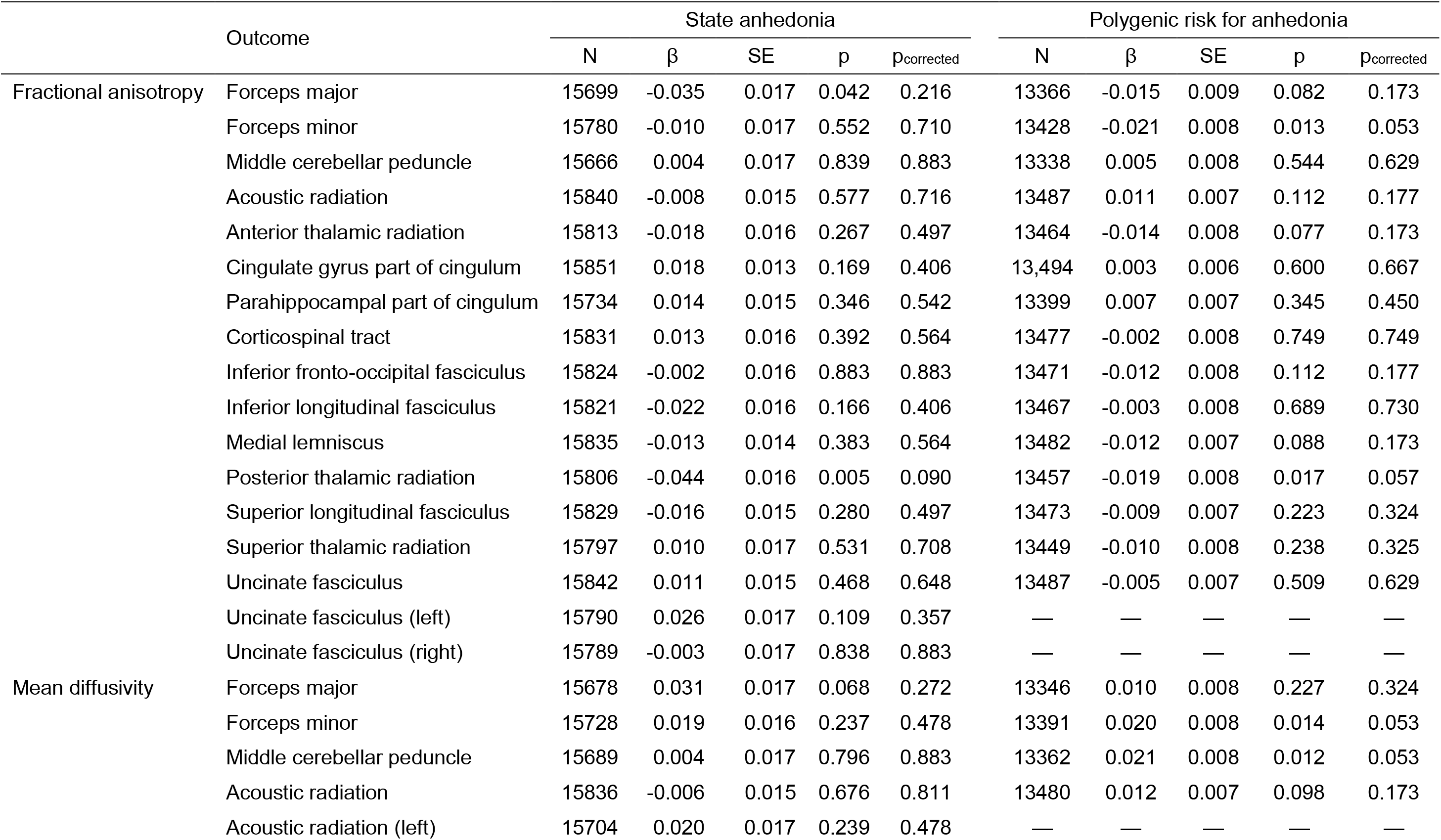
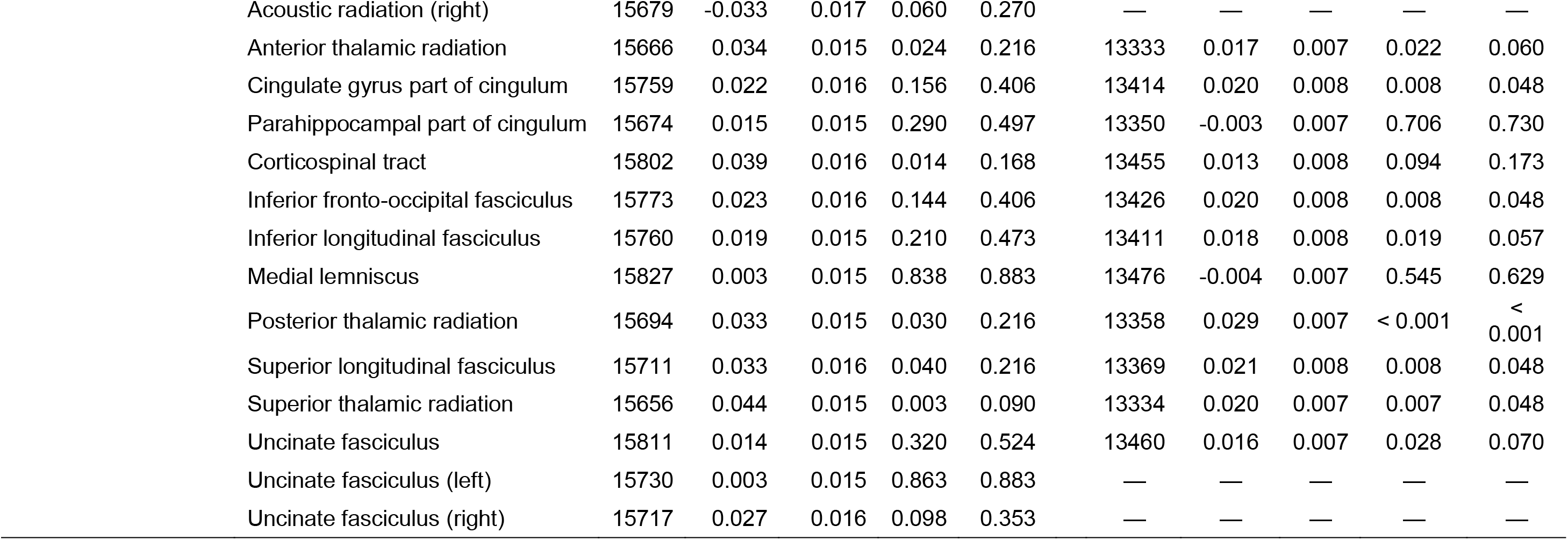
The associations between anhedonia, PRS-anhedonia and fractional anisotropy and mean diffusivity values of white matter tracts.

|  | Outcome | State anhedonia |  |  |  |  | Polygenic risk for anhedonia |  |  |  |  |
| --- | --- | --- | --- | --- | --- | --- | --- | --- | --- | --- | --- |
| | | N | $\beta$ | SE | p | p<sub>corrected</sub> | N | $\beta$ | SE | p | p<sub>corrected</sub> |
| Fractional anisotropy | Forceps major | 15699 | -0.035 | 0.017 | 0.042 | 0.216 | 13366 | -0.015 | 0.009 | 0.082 | 0.173 |
|  | Forceps minor | 15780 | -0.010 | 0.017 | 0.552 | 0.710 | 13428 | -0.021 | 0.008 | 0.013 | 0.053 |
|  | Middle cerebellar peduncle | 15666 | 0.004 | 0.017 | 0.839 | 0.883 | 13338 | 0.005 | 0.008 | 0.544 | 0.629 |
|  | Acoustic radiation | 15840 | -0.008 | 0.015 | 0.577 | 0.716 | 13487 | 0.011 | 0.007 | 0.112 | 0.177 |
|  | Anterior thalamic radiation | 15813 | -0.018 | 0.016 | 0.267 | 0.497 | 13464 | -0.014 | 0.008 | 0.077 | 0.173 |
|  | Cingulate gyrus part of cingulum | 15851 | 0.018 | 0.013 | 0.169 | 0.406 | 13,494 | 0.003 | 0.006 | 0.600 | 0.667 |
|  | Parahippocampal part of cingulum | 15734 | 0.014 | 0.015 | 0.346 | 0.542 | 13399 | 0.007 | 0.007 | 0.345 | 0.450 |
|  | Corticospinal tract | 15831 | 0.013 | 0.016 | 0.392 | 0.564 | 13477 | -0.002 | 0.008 | 0.749 | 0.749 |
|  | Inferior fronto-occipital fasciculus | 15824 | -0.002 | 0.016 | 0.883 | 0.883 | 13471 | -0.012 | 0.008 | 0.112 | 0.177 |
|  | Inferior longitudinal fasciculus | 15821 | -0.022 | 0.016 | 0.166 | 0.406 | 13467 | -0.003 | 0.008 | 0.689 | 0.730 |
|  | Medial lemniscus | 15835 | -0.013 | 0.014 | 0.383 | 0.564 | 13482 | -0.012 | 0.007 | 0.088 | 0.173 |
|  | Posterior thalamic radiation | 15806 | -0.044 | 0.016 | 0.005 | 0.090 | 13457 | -0.019 | 0.008 | 0.017 | 0.057 |
|  | Superior longitudinal fasciculus | 15829 | -0.016 | 0.015 | 0.280 | 0.497 | 13473 | -0.009 | 0.007 | 0.223 | 0.324 |
|  | Superior thalamic radiation | 15797 | 0.010 | 0.017 | 0.531 | 0.708 | 13449 | -0.010 | 0.008 | 0.238 | 0.325 |
|  | Uncinate fasciculus | 15842 | 0.011 | 0.015 | 0.468 | 0.648 | 13487 | -0.005 | 0.007 | 0.509 | 0.629 |
|  | Uncinate fasciculus (left) | 15790 | 0.026 | 0.017 | 0.109 | 0.357 | — | — | — | — | — |
|  | Uncinate fasciculus (right) | 15789 | -0.003 | 0.017 | 0.838 | 0.883 | — | — | — | — | — |
| Mean diffusivity | Forceps major | 15678 | 0.031 | 0.017 | 0.068 | 0.272 | 13346 | 0.010 | 0.008 | 0.227 | 0.324 |
|  | Forceps minor | 15728 | 0.019 | 0.016 | 0.237 | 0.478 | 13391 | 0.020 | 0.008 | 0.014 | 0.053 |
|  | Middle cerebellar peduncle | 15689 | 0.004 | 0.017 | 0.796 | 0.883 | 13362 | 0.021 | 0.008 | 0.012 | 0.053 |
|  | Acoustic radiation | 15836 | -0.006 | 0.015 | 0.676 | 0.811 | 13480 | 0.012 | 0.007 | 0.098 | 0.173 |
|  | Acoustic radiation (left) | 15704 | 0.020 | 0.017 | 0.239 | 0.478 | — | — | — | — | — |

|  |  |  |  |  |  |  |  |  |  |  |
| --- | --- | --- | --- | --- | --- | --- | --- | --- | --- | --- |
| Acoustic radiation (right) | 15679 | -0.033 | 0.017 | 0.060 | 0.270 | — | — | — | — | — |
| Anterior thalamic radiation | 15666 | 0.034 | 0.015 | 0.024 | 0.216 | 13333 | 0.017 | 0.007 | 0.022 | 0.060 |
| Cingulate gyrus part of cingulum | 15759 | 0.022 | 0.016 | 0.156 | 0.406 | 13414 | 0.020 | 0.008 | 0.008 | 0.048 |
| Parahippocampal part of cingulum | 15674 | 0.015 | 0.015 | 0.290 | 0.497 | 13350 | -0.003 | 0.007 | 0.706 | 0.730 |
| Corticospinal tract | 15802 | 0.039 | 0.016 | 0.014 | 0.168 | 13455 | 0.013 | 0.008 | 0.094 | 0.173 |
| Inferior fronto-occipital fasciculus | 15773 | 0.023 | 0.016 | 0.144 | 0.406 | 13426 | 0.020 | 0.008 | 0.008 | 0.048 |
| Inferior longitudinal fasciculus | 15760 | 0.019 | 0.015 | 0.210 | 0.473 | 13411 | 0.018 | 0.008 | 0.019 | 0.057 |
| Medial lemniscus | 15827 | 0.003 | 0.015 | 0.838 | 0.883 | 13476 | -0.004 | 0.007 | 0.545 | 0.629 |
| Posterior thalamic radiation | 15694 | 0.033 | 0.015 | 0.030 | 0.216 | 13358 | 0.029 | 0.007 | < 0.001 | <<br>0.001 |
| Superior longitudinal fasciculus | 15711 | 0.033 | 0.016 | 0.040 | 0.216 | 13369 | 0.021 | 0.008 | 0.008 | 0.048 |
| Superior thalamic radiation | 15656 | 0.044 | 0.015 | 0.003 | 0.090 | 13334 | 0.020 | 0.007 | 0.007 | 0.048 |
| Uncinate fasciculus | 15811 | 0.014 | 0.015 | 0.320 | 0.524 | 13460 | 0.016 | 0.007 | 0.028 | 0.070 |
| Uncinate fasciculus (left) | 15730 | 0.003 | 0.015 | 0.863 | 0.883 | — | — | — | — | — |
| Uncinate fasciculus (right) | 15717 | 0.027 | 0.016 | 0.098 | 0.353 | — | — | — | — | — |

### PRS-anhedonia and brain morphology

Analyses of whole-brain measures found that PRS-anhedonia was associated with lower total GMV (β = −0.015, p_corrected_ < 0.001; Table 2) and higher total WMV (β = 0.013, p_corrected_ < 0.001). There was no significant interaction between PRS-anhedonia and hemisphere on subcortical volumes, as well as regional CT (Table S6). We found significantly negative associations between PRS-anhedonia and CT in the insula cortex (β = −0.029, p_corrected_ < 0.001; Table 2; Fig. 1B), parahippocampal cortex (PHC; β = − 0.031, p_corrected_ < 0.001) and superior temporal gyrus (STG; β = −0.023, p_corrected_ = 0.008).

### PRS-anhedonia and white matter integrity

We found that none of the measures of white matter integrity demonstrated significant interaction of hemisphere (Table S7), and none of the associations between PRS-anhedonia and FA values survived correction for multiple testing (Table 3). However, PRS-anhedonia was associated with higher in MD within the SLF (β = 0.021, p_corrected_ = 0.048; Table 3), the cingulate gyrus part of cingulum (β = 0.020, p_corrected_ = 0.048), the inferior fronto-occipital fasciculus (β = 0.020, p_corrected_ = 0.048), the superior thalamic radiation (β = 0.020, p_corrected_ = 0.048) and the posterior thalamic radiation (β = 0.029, p_corrected_ < 0.001).

### Sensitivity analyses

Analyses for anhedonia as a dichotomous variable found similar results, except that CT in the paracentral gyrus became marginally significant (Table S8). In addition, we found that patients with depression/post-natal depression had more severe anhedonia (t = −26.802, p < 0.001) compared to healthy participants, and that there was no significant interaction between anhedonia and depression/post-natal depression (Table S9), consistent with previous reports (30). As shown in Fig. S3, the significant outcomes from the main analyses showed a similar pattern in both healthy people and in those with a history of depression.

Furthermore, when Townsend social deprivation index, education qualification, BMI, current tobacco smoking status and alcohol intake frequency were included in the model, the association between anhedonia and brain measures reported previously showed a similar pattern (Table S10). Re-analyses for PRS-anhedonia including additional covariates also found significant associations with smaller brain volumes and thinner cortices (Table S11), together with poor white matter integrity (Table S12).

### Mediation analyses

Firstly, we examined the potential mediation effect of brain measures and found none of them significantly mediated the effect of PRS-anhedonia on state anhedonia. Conversely, in the model with each brain measure set as an outcome, we found that state anhedonia significantly mediated the effect of PRS-anhedonia on total GMV (indirect effect: β = −0.0004, p_corrected_ = 0.049), volume of the thalamus (indirect effect: β = −0.001, p_corrected_ = 0.040) and NAcc (indirect effect: β = −0.0015, p_corrected_ = 0.030), CT in the pars opercularis (indirect effect: β = −0.0012, p_corrected_ = 0.049), precentral (indirect effect: β = −0.0011, p_corrected_ = 0.049) and rostral anterior cingulate (indirect effect: β = −0.0012, p_corrected_ = 0.049), as well as. FA in the superior thalamic radiation (indirect effect: β = 0.0011, p_corrected_ = 0.049). Moreover, depression played a mediating role in the relationship between PRS-anhedonia and MD in the SLF (indirect effect: β = 0.0009, p_corrected_ = 0.049), the cingulate gyrus part of cingulum (indirect effect: β = 0.0011, p_corrected_ = 0.040), the inferior fronto-occipital fasciculus (indirect effect: β = 0.0008, p_corrected_ = 0.049) and the superior thalamic radiation (indirect effect: β = 0.0008, p_corrected_ = 0.049). More detail on these mediation analyses is provided within supplementary Data.

## Discussion

Overall, we found that the phenotype of state anhedonia and a measure of polygenic risk for anhedonia were both associated with differences in brain structure. State anhedonia was associated with smaller volumes in the thalamus and NAcc, and with lower cortical thickness in the paracentral gyrus, pars opercularis and rostral ACC. No significant associations between state anhedonia and white matter integrity indices were identified. PRS-anhedonia analyses showed that higher polygenic risk was associated with a range of brain morphology differences, including lower total GMV, higher total WMV, thinner PHC, STG and insula cortex, plus higher MD in the SLF, the cingulate gyrus part of cingulum, the inferior fronto-occipital fasciculus, the superior thalamic radiation and the posterior thalamic radiation. Further, although we could not determine the directional or causal relationship of these associations, our mediation analyses suggested that state anhedonia/depression might mediate the effect of PRS-anhedonia on brain structure.

### State anhedonia and brain structures

Consistent with previous studies, state anhedonia was found to be associated with loss of grey matter in the thalamus, NAcc and rostral ACC, confirming a fronto-striatal reward circuit abnormality (6-8). Previous studies have found anhedonia to be associated with reduced volume or CT in these regions within non-clinical populations (14, 15), as well as in patients with schizophrenia and MDD (10, 31-33). In addition, we found a negative association between state anhedonia and CT in the paracentral cortex and pars opercularis, consistent with previous studies in patients with schizophrenia (9, 10, 34, 35), corresponding to their potential roles in reward anticipation to some extent (36, 37). Neuroanatomical alterations in these reward-related brain regions may lead to functional deficits in motivation and goal-directed behavior, thereby giving rise to loss of the capacity to experience pleasure.

### PRS-anhedonia and brain structures

Our analyses for PRS-anhedonia found associations with reduced total GMV and increased total WMV, together with reduced CT in the PHC, STG and insula cortex. Previous findings on total GMV and total WMV are inconsistent. Van Scheltinga *et al*. (38) reported reduced total brain volume and total WMV in relation to PRS for schizophrenia and no significant association with GMV. Other studies have found no associations between PRS for schizophrenia/MDD and GMV and or WMV (39-41). However, it is notable that these studies had relatively small sample sizes (ranging from 152 to 1470), perhaps contributing to a lack of power to detect associations.

Regarding subcortical volume measures, we found no significant associations with PRS-anhedonia, consistent in accord with previous studies of PRS for schizophrenia and MDD (39, 41, 42). For cortical regions, our prior work (4) on GMV also reported negative associations in the insular and temporal cortex, in addition to the findings on CT described in the current study. Results from PRS for schizophrenia also indicated reduced insular CT (43). Evidence from functional MRI studies has found that PRS for schizophrenia was associated with brain activity in the insula and superior temporal gyrus during emotion-processing (44). The cross-disorder PRS of five psychiatric disorders also reported altered functional connectivity involving the bilateral <u>insula</u> (45). Moreover, studies of CT in patients with schizophrenia and MDD have reported widespread thinning of the cortex, including PHC, STG and insula cortex (35, 46). A meta-analysis for emotion/reward tasks in relation to anhedonia also observed associations with altered activation in widespread regions including PHC, STG and insula cortex in healthy controls and patients with schizophrenia or MDD (47).

### PRS-anhedonia and white matter integrity

We found associations between PRS-anhedonia and higher MD in the SLF, the cingulate gyrus part of cingulum, the inferior fronto-occipital fasciculus, the superior thalamic radiation and the posterior thalamic radiation, in line with previously reported results by Ward *et al*. (4) from our group in 2019. This study was conducted in a subsample of participants included in Ward *et al*. (4) because we excluded those without data of state anhedonia. Given that there are potential shared genetic components between white matter integrity and MDD and schizophrenia (48-50) - and genetic correlations between anhedonia, MDD and schizophrenia (4, 23) - it is perhaps not surprising that we observed associations between polygenic risk for anhedonia and white matter indices. In over 9,000 participants from UK Biobank, Shen *et al*. (51) recently found PRS for MDD was associated with white matter integrity of several tracts, such as the SLF, the cingulate gyrus part of cingulum, the inferior fronto-occipital fasciculus and superior thalamic radiation, which were associated with PRS-anhedonia in our study. Previous studies on PRS for MDD have also reported associations with decreased white matter integrity, most notably in the SLF, cingulum and thalamic radiations (52-54), although null findings have also been reported with respect to polygenic risk for MDD or schizophrenia (39, 55). Together with our findings of worse white matter integrity in relation to state anhedonia (despite not statistically significant), white matter structural abnormalities associated with PRS-anhedonia could be a biomarker of vulnerability to anhedonia and psychiatric disorders.

### Potential mediation candidates

Using mediation analyses, we did not identify specific brain regions or white matter tracts that could mediate the relationship between PRS-anhedonia and state anhedonia, while we found that state anhedonia mediated the link between PRS-anhedonia and volume in the NAcc. To the best of our knowledge, no study has examined mediation candidates of polygenic risk for anhedonia. With regard to polygenic risk for MDD, Shen *et al*. (51) found that current depressive symptoms mediated the effect of depression-PRS on global MD, MD in thalamic radiations and SLF, and that MD in ATR mediated the effect of depression-PRS on current depressive symptoms, as well as a significant causal effect of depression on lower white matter integrity. This indicates that the relationship between psychosis and brain phenotypes may not be one-way causality and there may be complex interactions between psychosis and the brain. More studies are needed to clarify the associations between psychiatric disorders and the brain. We will also explore the directional or causal relationship between anhedonia and brain measures and identify putative neural mediators in the future.

### Strengths and Limitations

To our knowledge, this is the largest study to test for associations between anhedonia and brain measures within a single, non-clinical, population-based sample, and the first study to examine associations between anhedonia, PRS-anhedonia, and cortical thickness. This study excluded all depression and schizophrenia cases and those with other mental illness, thus the associations we detected were not driven by confounding factors such as mental illness or use of antipsychotic medication, but rather by the genetic risk captured by the PRS-anhedonia.

Despite these strengths, there are some important potential limitations. Firstly, this sample is relatively large compared to previous imaging PRS studies (e.g. 39, 41, 43), with 17,492 individuals, but may still be underpowered to detect a robust mediation effect or significant mediation of brain structures, highlighting the need for much larger imaging samples in the future. Secondly, we note that participants in this study were aged from 45 to 80, which means that cumulative environmental risk may contribute to the associations we found. Thirdly, considering that anhedonia may also manifest in childhood, before the onset of psychiatric disorders (5, 56), further consideration of how polygenic risk for anhedonia may influence child or adolescent brain development and brain changes over longer developmental periods will be important to further understand the nature of the relationship between anhedonia and brain structure. Finally, another potential limitation concerns the measurement of anhedonia as a single item self-report: a dimensional measure of anhedonia, such as the Chapman Physical and Social Anhedonia Scale (57) might prove to be a more informative phenotype.

## Conclusion

In summary, we found that the symptom of anhedonia and genetic risk for anhedonia were both associated with brain structural differences within a large population-based sample of 17,492 people. This included differences in total grey/white matter volume, thalamus, NAcc, the paracentral gyrus, pars opercularis, rostral ACC, PHC, STG and insula, as well as several white matter tracts. Importantly, this is the first large-scale investigation of how cortical thickness may be associated with anhedonia. Overall, our findings suggest that reward-related brain structural differences are associated with anhedonia and its genetic risk and highlight potential neuroanatomical markers of risk for psychopathology across a range of psychiatric disorders.

## Data Availability

All the data used in this manuscript comes from UK Biobank.

https://www.ukbiobank.ac.uk/

## Acknowledgments

XZ acknowledges financial support from China Scholarship Council. RJS is funded by UKRI Innovation-HDR-UK Fellowship(MR/S003061/1). DJS acknowledges support from a Lister Institute Prize Fellowship and an MRC Mental Health Data Pathfinder Award. JW is funded by DJS Lister Institute Prize Fellowship. LML is supported by the JMAS Sim Fellowship.

